# ELABELA as a Potential Diagnostic Biomarker and Therapeutic Target of Atherosclerosis

**DOI:** 10.1101/2024.11.07.24316940

**Authors:** Le Tang, Xiaoli Yi, Huiru Yang, Shanshan Song, Wenting Tan, Jianhua Xiong, Chunju Liu, Yifeng Zhang, Mulan Wang, Mengzhi Zhu, Lixiang Zheng, Jun Yu, Chuanming Xu

**Affiliations:** Translational Medicine Centre, Jiangxi University of Chinese Medicine, Nanchang 330004, China; Department of Cardiology, Affiliated Hospital of Jiangxi University of Chinese Medicine, Nanchang 330006, China; Department of Clinical Laboratory, Affiliated Hospital of Jiangxi University of Chinese Medicine, Nanchang 330006, China; College of Traditional Chinese Medicine, Jiangxi University of Chinese Medicine, Nanchang 330004, China; Center for Metabolic Disease Research and Department of Cardiovascular Sciences, Lewis Katz School of Medicine, Temple University, Philadelphia, PA 19140, USA

**Author notes:** Address correspondence to: Jun Yu, Center for Metabolic Disease Research and Department of Cardiovascular Sciences, Lewis Katz School of Medicine, Temple University, Philadelphia, PA 19140, USA.,; Chuanming Xu, Ph.D., Translational Medicine Centre, Jiangxi University of Chinese Medicine, Nanchang 330004, Jiangxi, China.

**Keywords:** Atherosclerosis, ELABELA, Macrophage, Diagnosis, Therapy, renin-angiotensin system

## Abstract

Atherosclerosis (AS) is a progressive arterial disease characterized by chronic inflammation and plaque formation in blood vessel walls. ELABELA, an endogenous ligand for the G protein-coupled receptor APJ (apelin peptide jejunum, apelin receptor), has multiple pharmacological activities for protecting the cardiovascular system. This study aimed to determine the potential anti-atherosclerotic effect of ELABELA and reveal the underlying mechanisms. Plasma ELABELA levels were significantly reduced and negatively correlated with plasma MMP2 and MMP9 levels in AS patients and high-fat diet-induced atherosclerotic *ApoE*^−/−^ mice. Plasma ELABELA levels exhibited a potential diagnostic value for AS patients. Application of ELABELA-21 (ELA-21) significantly decreased atherosclerotic plaque area and inflammation in the aortas from the *ApoE*^-/-^ mice. ELA-21 administration modulated the balance between M1 and M2 macrophages in the abdominal cavity and aorta roots toward a more anti-inflammatory status, accompanied by reduced MMP2, MMP9, and PRR and enhanced APJ, ACE, and ACE2 protein expression in plaques within aortic roots and decreased plasma sPRR levels. *In vitro*, ELA-21 effectively suppressed oxidized-low-density lipoprotein-induced foam cell formation and LPS/IFN-γ-induced M1 polarization in THP-1 cells. Interestingly, the anti-inflammatory effect of ELA-21 was further enhanced by APJ inhibitor ML221, accompanied by elevated *ACE* and *ATP6AP2* and reduced *ACE2* mRNA levels. Collectively, our data highlighted the diagnostic and therapeutic potential of ELABELA on AS. ELA-21 protects against AS by inhibiting atherosclerotic plaque formation and promoting a more stable plaque phenotype, possibly via restoring the M1/M2 macrophage balance, enhancing macrophage ACE and ACE2 expression, and inhibiting the PRR system. ELABELA may be a novel biomarker and candidate therapeutic target for treating AS.

## Introduction

Atherosclerosis (AS), a chronic inflammatory disease, is not only the leading cause of the morbidity and mortality of cardiovascular diseases globally but also the primary pathological basis of most cardiovascular diseases^[1]^. AS is initialed by endothelial dysfunction, followed by the adhesion of monocytes, monocytes differentiate into macrophages under the stimulation of macrophage-stimulating factor and other cytokines, and the formation of foam cells after the engulfment of oxidized low-density lipoprotein (ox-LDL) by macrophages, the release of contents from the dead foam cells, and gradual aggregation of vascular smooth muscle cells (VSMCs), collagen, and foam cells, eventually forming atherosclerotic plaque^[2]^. Currently, the first-line drugs widely used in clinical practice for AS are mainly lipid-lowering drugs, such as statins [inhibiting HMG-CoA (3-hydroxy-3-methyl glutaryl coenzyme A) reductase], beta drugs, and preprotein converting enzyme subtilisin 9 (PCSK9) inhibitors ^[3]^. In addition, antiplatelet drugs, such as aspirin^[4]^ and clopidogrel ^[5]^, and anti-inflammatory drugs, such as renin-angiotensin system (RAS) inhibitors ^[6]^, are also commonly used in clinical practice for AS. However, although the above drugs have slowed down AS progression to a certain extent, their efficacy in reducing the mortality of cardiovascular diseases is only about 30%. They are accompanied by potential adverse reactions. For instance, ezetimibe can cause respiratory infection, muscle pain, and joint pain, and RAS inhibitors can cause hyperkalemia. Therefore, novel, innovative, and safer pharmacological interventions and anti-atherosclerotic drugs are needed to ameliorate atherosclerotic cardiovascular diseases and, thus, mortality worldwide.

The apelineargic system, an important endocrine system in the body, consists of two endogenous peptide ligands, apelin (encoded by *Apln*) and ELABELA (encoded by *Apela* and also known as Toddler), as well as the G protein-coupled receptor APJ (Apelin receptor, encoded by *Aplnr*)^[7]^. In human atherosclerotic plaques, apelin and APJ are co-localized with α-SMA in VSMCs and with CD68 in macrophages ^[8]^. Although upregulated apelin and APJ levels were observed in human atherosclerotic coronary artery^[8]^, increasing studies have reported decreased plasma apelin levels in patients with acute coronary syndrome^[9, 10]^, rheumatoid arthritis^[11]^, and symptomatic intracranial atherosclerotic stenosis^[12]^. In particular, plasma apelin levels were positively associated with plasma matrix metallopeptidase 2 (MMP2) levels but negatively correlated with plasma MMP9 levels in patients with rheumatoid arthritis^[11]^, implying a positive correlation between plasma apelin levels and atherosclerotic plaque stability. This notion was supported by the results observed in patients with acute coronary syndrome that plasma apelin levels were significantly lower in patients with the ruptured plaque than in those with the non-ruptured plaque and inversely associated with plaque cross-sectional area (CSA) but positively related with external elastic membrane CSA^[9]^. What’s more, apelin can not only inhibit the formation of foam cells by activating cell autophagy and inhibiting lipid accumulation in macrophages, but also promote M2 polarization of macrophages, thereby inhibiting the release of pro-inflammatory factors^[13–15]^, implying the potential therapeutic action of apelin on AS. Indeed, apelin-13 has been reported to inhibit AngII-induced AS in *ApoE*^−/−^ mice by increasing nitric oxide bioavailability^[16]^. Similarly, apelin-13 also significantly enhanced the stability of atherosclerotic plaque by increasing the collagen content in the plaque and reducing the expression of MMP-9, the infiltration of inflammatory cells (neutrophils and macrophages), and the levels of reactive oxygen species in the plaque, without affecting the lesion size in high-fat diet (HFD)-induced atherosclerotic *ApoE*^−/−^ mice^[17]^. Overall, these results suggest the important clinical value of apelin in the diagnosis and therapy of AS. Of note, Hashimoto *et al*. have reported that systemic APJ knockout significantly reduced atherosclerotic lesions by inhibiting oxidative stress in VSMCs and cell proliferation, without affecting cholesterol levels in high-cholesterol-fed *ApoE*^-/-^ mice^[18]^. These results may imply that the regulatory effect of APJ on AS seems to be opposite to that of apelin, or the anti-atherosclerotic effects of apelin may be independent of APJ.

Similar to the bioeffects of apelin, ELABELA also displays significant protective actions on the cardiovascular system^[7]^. A study has demonstrated a decreased circulating ELABELA level in hypertensive patients with an increased carotid intima-media thickness^[19]^, implying a potential negative correlation between circulating ELABELA and subclinical AS. However, the clinical significance of ELABELA in AS patients and the impact of ELABELA on AS progression remain unclear. The aims of this study are: 1) to determine plasma ELABELA levels in AS patients and assess the correlation between plasma ELABELA and AS severity and 2) to evaluate the potential therapeutic effect of ELA-21 on AS in HFD-fed *ApoE*^−/−^ mice.

## Methods

### Study Population

The study protocol was approved by the Ethics Committee of the Affiliated Hospital of the Jiangxi University of Chinese Medicine (JZFYLL20230208002) and performed under the declaration of Helsinki-Ethical principles. All participants provided written informed consent and clinical characteristics after fully explaining the purpose of the study and the potential risk involved. The inclusion and exclusion criteria for enrolled AS patients and the collection method of plasma samples were previously described in detail^[20]^. Briefly, the diagnosis of subclinical AS was established after evaluating the carotid intima-media thickness and plaque area according to the bilateral carotid ultrasonography examination; patients with heart diseases (heart failure, rheumatic heart disease, valvular heart disease, and cardiomyopathy), hepatic failure, kidney disease (chronic and acute kidney injury), hyperthyroidism, malignancy, pulmonary embolism, and other inflammatory diseases were excluded. Blood samples were collected from 236 patients/participants between December 2022 and December 2023 at the Department of Cardiology, Affiliated Hospital of Jiangxi University of Chinese Medicine. All the fasting blood samples were collected from a peripheral vein of all patients within 24-h of admission and immediately centrifuged for 5 min at 4 °C and 3,000 rpm to separate plasma, which was immediately stored at −80 °C until use. None of the patients/participants received the optimized treatment before collecting blood samples. The participants were divided into Non-AS and AS groups. All the laboratory assessments, except plasma ELABELA and apelin levels, were conducted in the clinical laboratory center of the Affiliated Hospital of the Jiangxi University of Chinese Medicine according to the standard protocols.

### High-fat-induced atherosclerosis in *ApoE*^-/-^ mice

Male and female *ApoE*^−/−^ mice (6-8-week-old) were purchased from Nanjing Biomedical Research Institute of Nanjing University, Nanjing, China. Mice were all given free access to tap water and the standard diet. Mice were housed in individually ventilated cages (temperature 20-26 °C, humidity 40-70%) with a 12:12-h light-dark cycle. The Animal Care and Use Committee approved the animal protocols at the Jiangxi University of Chinese Medicine (No. JZLLSC20230254), and all the processes are in strict accordance with the National Institutes of Health (NIH) Guide for the Care and Use of Animals in laboratory experiments and ARRIVE guidelines. All mice were received HFD (42% fat) feeding for 12 weeks and then randomly divided into two groups: mice in the HFD group were received intraperitoneal injection of vehicle (0.9% physiological saline), mice in the HFD+ ELABELA-21 (ELA-21) group were received intraperitoneal injection of ELA-21 (LRKHNCLQRRCMPLHSRVPFP, >98% purity, GenScript) in 0.9% physiological saline (1 mg/kg/day) for additional 4 weeks. At the end of the experiments, mice were anesthetized by isoflurane, and then plasma, peritoneal macrophages, full-length aorta, and the aortic root were harvested for further analysis. The technology roadmap of this study is shown in **Fig. 3A**.

### En face Oil-Red-O staining of full-length aortas

The fat-stripped full-length aortas of *ApoE*^−/−^ mice were fixed at 4 ℃ in 4% paraformaldehyde/0.1 M phosphate salt buffer and dehydrated with 30% sucrose gradient. The aortas were longitudinally cut open to fully expose the inner surface, followed by two equilibration steps in 60% isopropanol for 5 min each and stained with 0.3% Oil-Red-O staining solution at room temperature for 20 min in the dark. The aortas were differentiated with 60% isopropanol 2∼3 times and rinsed with distilled water to terminate the differentiation and remove the excess dye. Images were captured and the atherosclerotic lesion areas were measured using Image-Pro Plus version 6.0 software.

### Histological examination and immunofluorescence staining of aortic root lesions

The aortic roots were fixed overnight at 4 ℃ in 4% paraformaldehyde/0.1 M phosphate salt buffer and dehydrated with 30% sucrose gradient. The samples were embedded in an optimal cutting temperature compound and frozen using liquid nitrogen. Ten-micrometer cryosections were obtained from the aortic root to the apex. Slides were used for H&E, Oil-Red-O, and Masson staining for histological examination according to the manufacturers’ instructions. Antibodies used for immunofluorescence staining were shown in **Table S1**. Images were captured using a Leica DMI4000B fluorescence microscope (Wetzlar, Germany). The quantification of the sizes of both atherosclerotic plaque and collagen fibers was analyzed using Image-Pro Plus version 6.0 software, and the average fluorescence intensity was quantified by calculating the total area of fluorescence intensity relative to the plaque area.

### Plasma biochemical analysis

Plasma collected was instantaneously centrifuged at 3000 rpm for 10 min at 4 ℃. The plasma concentrations of ELABELA (S-1508, Peninsula Laboratories International, Inc. USA), Apelin (For human sample: E01T0015, Bluegene Tech Inc., Shanghai, China; for mouse sample: EKE-057-15, Phoenix Pharmaceuticals Inc., California, USA), and sPRR (27782, Immuno-Biological Laboratories, Gunma, Japan) were evaluated by commercial ELISA kits according to the manufacturer’s instructions, respectively. ELISA measurements of plasma MMP2 (HM10737 for human, MU30640 for mouse,), MMP9 (HM10095 for human, MU30613 for mouse), IL-1β (MU30369), IL-6 (MU30044), TNF-α (MU30030), and IL-10 (MU30055) were performed following the manufacturer’s instructions (Bioswamp, Wuhan, China).

### Transcriptome Sequencing and Data Processing

Aortic samples in HFD and HFD + ELA-21 groups (n = 3 for each group) were immediately harvested for total RNA extraction and RNA quality was estimated to construct the transcriptomics sequencing library. Transcriptome sequencing and analysis were performed by Novogene Biotechnology (Beijing, China) using an Illumina Novaseq platform. Reads from all samples were mapped to the GRCm39 reference genome using the HISAT2 package (HISAT 2.2.1). Differential expression analysis between the two groups was conducted using the DESeq2 software (v3.19). In this work, |log2FoldChange| ≥1.5 and Padj <0.05 were set as the threshold for the definition of differentially expressed genes (DEGs). All DEGs were initially mapped to GO terms in the Gene Ontology database to identify Top10 enriched GO terms among the DEGs with a significance threshold of p<0.05. Pathway enrichment analysis was conducted using the KEGG database to identify Top10 enriched signal transduction pathways among the DEGs with an adjusted p-value<0.05.

### Cell culture

The THP-1 and RAW264.7 cells were cultured in RPMI 1640 medium containing 10% FBS at 37 ℃ with 5% CO_2_. The THP-1 and RAW264.7 cells were treated with 100 ng/mL PMA for 24 h and then pretreated with ELA-21 (0.01, 0.1, 1 or 5 μM) for 12 h, followed by LPS/IFN-γ or ox-LDL treatment for 24 h. The expression of inflammatory cytokines or the formation of foam cells was detected.

### Quantitative Reverse Transcriptase PCR (RT-qPCR)

Total RNA was extracted from aortic samples, peritoneal macrophages, THP-1 cells, and RAW264.7 cells using Trizol reagent (15596018, Life Technologies, Carlsbad, CA, USA) and was reversed transcribed to cDNA using the HiScript Q RT SuperMix (R122, Vazyme, Nanjing, China). Real-time qPCR was performed using the Hieff® qPCR SYBR Green Master Mix reagent (11201ES, Yeasen, Shanghai, China) and specific primer (**Table S2**) in the Light Cycler® 96 System (Roche, Ricardo Rojas, Argentina). Relative mRNA expression of each gene was shown as a relative value normalized by *18s* or *GAPDH*.

### Statistical analysis

Continuous data in the cohort study were expressed as mean ± standard deviation (SD) for normally distributed data, median and interquartile range for non-normally distributed data, and categorical variables as number and percentage. Data in the animal study were summarized as means ± SD. Unpaired Student’s t-test performed statistical analysis for two comparisons using IBM SPSS 19 software. Spearman correlation analysis was used to correlate plasma ELABELA or Apelin levels and study variables. The diagnostic value of plasma ELABELA and Apelin was assessed by determining the area under the receiver operating characteristic (ROC) curves (AUC) using IBM SPSS 19 software. All tests were two-sided, and *p* < 0.05 was considered statistically significant.

## Results

### Plasma ELABELA and apelin levels in patients with and without AS

The baseline characteristics of enrolled non-AS patients (n = 63, age 58.8±11.7 years old) and AS patients (n = 173, age 67.1±11.0 years old, *p*<0.001 vs. non-AS group) have been described in detail in our previous studies^[20]^. Briefly, there were no significant differences in gender, body mass index (BMI), comorbidities, or blood pressure between the two groups. Laboratory examinations revealed that plasma BNP, urea nitrogen, and triglyceride levels were slightly higher, and plasma MMP2 (4.4±1.8 vs. 3.5±1.9 ng/ml, p<0.001) and MMP9 (2.7±1.2 vs. 1.7±0.9 ng/ml, p<0.001) levels were significantly higher in the AS group than those in the non-AS group. In contrast, other parameters, including plasma creatine, uric acid, LDL-c, HDL-c, and total cholesterol, were not different between the two groups. According to the echocardiographic data, the left and right carotid maximum intima-media thickness (IMT) were significantly increased in AS group compared to those in non-AS group (*p*<0.001), while other parameters reflecting cardiac function including the left ventricular ejection fraction (LVEF) was lower and the left atrial diameter (LAD), left ventricular end-diastolic diameter (LVEDd), left ventricular end-systolic diameter (LVEDs), left ventricular posterior wall (LVPW) thickness, interventricular septum thickness (IVST), and right ventricular internal dimension diameter (RVIDd) were not significantly different between the two groups. The levels of plasma ELABELA in AS patients were significantly lower than those in non-AS groups (9.4±3.7 *vs*. 14.9±3.3 ng/ml, *p*<0.001, **Figure 1A**). The mean plasma Apelin (34.3±31.1 *vs*. 57.9±56.0 ng/ml, *p*<0.001, **Figure 1B**) levels of the AS groups were also significantly lower than those in the non-AS group.

**Figure 1.**
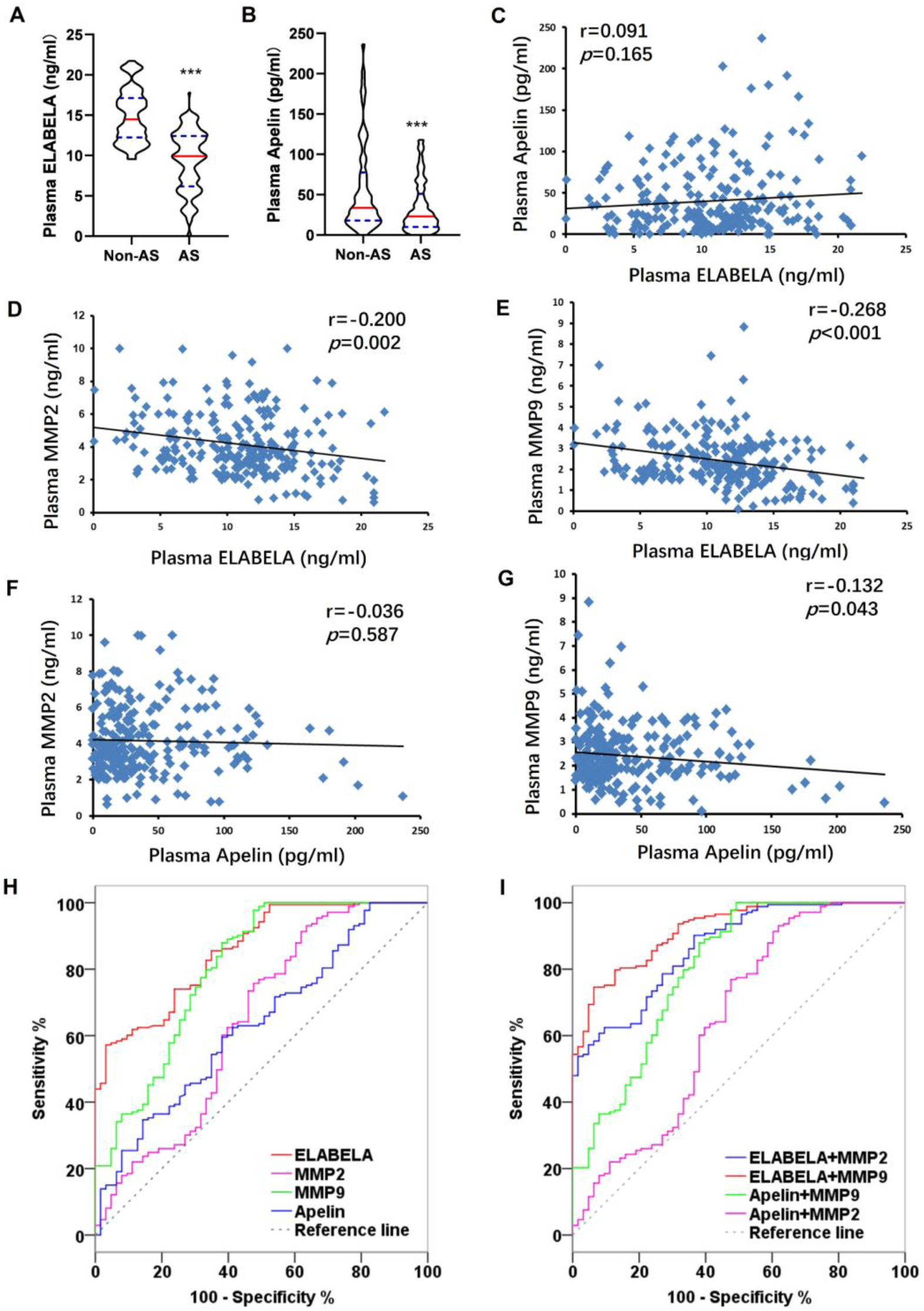
Plasma ELABELA and apelin levels in patients with atherosclerosis (AS). (A and B) plasma ELABELA (A) and apelin (B) levels assessed by ELISA assays. n=63 for the non-AS group and n=173 for the AS group. \*\*\**P*<0.001 *vs*. Non-AS. (C) The correlation between plasma ELABELA and apelin. (D and E) The correlation between plasma ELABELA and matrix metalloproteinase 2 (MMP2) (D) and MMP9 (E). (F and G) The correlation between plasma apelin and MMP2 (F) and MMP9 (G). (H and I) Receiver Operating Characteristic (ROC) curves of plasma ELABELA, apelin, MMP2, and MMP9 levels (H) as well as their combinations (I) for AS. AUC, area under curve; CI, confidence interval.

To estimate the diagnostic values of ELABELA, Apelin, MMP2/9, and their combinations for AS, the AUC was analyzed for data from all non-AS and AS patients (**Figure 1H-I, Table S3**), followed by a pairwise comparison of ROC curves via the DeLong test (**Table S4**). When a single diagnosis was used, ELABELA had the highest diagnostic value (AUC 0.855±0.025, 95% CI 0.803-0.897, sensitivity 56.65%, specificity 96.83%), followed by MMP9 (AUC 0.803±0.036, 95% CI 0.746-0.851, sensitivity 97.69%, specificity 52.38%), suggesting a potential diagnostic value of plasma ELABELA levels for AS with moderate sensitivity and high specificity. When multiple joint diagnoses were performed, ELABELA combined with MMP9 had the highest diagnostic value (AUC 0.919±0.018, 95% CI 0.877-0.951, sensitivity 74.57%, specificity 93.65%), followed by ELABELA+MMP2 (AUC 0.866±0.025, 95% CI 0.816-0.907, sensitivity 90.17%, specificity 63.49%), and apelin+MMP9 (AUC 0.802±0.036, 95% CI 0.745-0.851, sensitivity 100.00%, specificity 50.79%). Thus, a combined assessment of ELABELA and MMP2/MMP9 may be a good choice to increase the accuracy of the diagnosis of AS.

We analyzed the correlation between plasma ELABELA or Apelin and study variables in all subjects (**Table S5**). Age (r=-0.257, p<0.001), plasma MMP2 levels (r=-0.221, p=0.001, **Figure 1D**), plasma MMP9 levels (r=-0.280, p<0.001, **Figure 1E**), LAD (r=-0.495, p<0.001), LVEDs (r=-0.490, p<0.001), LVPW (r=-0.172, p=0.008), IVST (r=-0.538, p<0.001), left carotid maximum IMT (r=-0.307, p<0.001), right carotid maximum IMT (r=-0.284, p<0.001), left carotid plaque area (r=-0.330, p<0.001), and left carotid plaque area (r=-0.311, p<0.001) were negatively related to plasma ELABELA levels. Heart rate (r=0.182, p=0.005), plasma HDL-c levels (r=0.146, p=0.025), and LVEDd (r=0.560, p<0.001) were positively correlated to plasma ELABELA levels. In contrast, plasma MMP9 levels (r=-0.132, p=0.043, **Figure 1G**), LAD (r=-0.232, p<0.001), LVEDs (r=-0.223, p=0.001), IVST (r=-0.243, p<0.001), left carotid maximum IMT (r=-0.317, p<0.001), right carotid maximum IMT (r=-0.245, p<0.001), and left carotid plaque area (r=-0.146, p=0.024) were negatively related to plasma apelin levels. Plasma triglyceride levels (r=0.147, p=0.024) and LVEDd (r=0.282, p<0.001) were positively correlated to plasma Apelin levels. Multiple linear regression analyses in all the participants were further performed to determine the relationship between plasma ELABELA levels and plasma apelin levels and clinical characteristics associated with AS (**Table S6**). We found a significant association between plasma ELABELA levels and heart rate (β=0.109, t=2.877, p=0.004), LAD (β=0.217, t=3.392, p=0.001), left carotid maximum IMT (β=7.980, t=0.910, p=0.046), and right carotid maximum IMT (β=-9.535, t=-1.143, p=0.014), while plasma Apelin levels were only associated with left carotid maximum IMT (β=7.980, t=0.910, p=0.046) and right carotid maximum IMT (β=-9.535, t=-1.143, p=0.014).

### Plasma ELABELA and apelin levels in *ApoE*^−/−^ mice fed a HFD

To further assess the status of plasma ELABELA and apelin levels in the setting of AS, the *ApoE*^-/-^ mice were fed an HFD (42% fat) for 16 weeks to induce AS, and then the levels of plasma ELABELA and apelin were examined in these mice. After HFD feeding, both the levels of plasma ELABELA (5.7±3.6 *vs*. 17.3±3.2 ng/ml, *p*<0.001, **Figure 2A**) and apelin (3.3±0.9 *vs*. 5.0±0.6 ng/ml, *p*<0.001, **Figure 2B**) were significantly reduced. A significant upregulation of plasma MMP2 (9.7±1.1 *vs*. 6.3±1.1 ng/ml, *p*<0.001) and MMP9 (4.7±0.9 *vs*. 2.3±0.4 ng/ml, *p*<0.001) levels were observed in these mice^[20]^. We also analyzed the correlation between plasma ELABELA, apelin, and plasma MMPs in HFD-induced atherosclerotic mice. Plasma ELABELA levels were negatively correlated with plasma MMP2 (r=-0.647, *p*<0.001) (**Figure 2D**) and MMP9 levels (r=-0.627, *p*<0.001) (**Figure 2E**) in these mice. A similar negative correlation between plasma apelin levels and plasma MMP2 (r=-0.602, *p*<0.001) (**Figure 2F**) and MMP9 levels (r=-0.619, *p*<0.001) (**Figure 2G**) was also observed.

**Figure 2.**
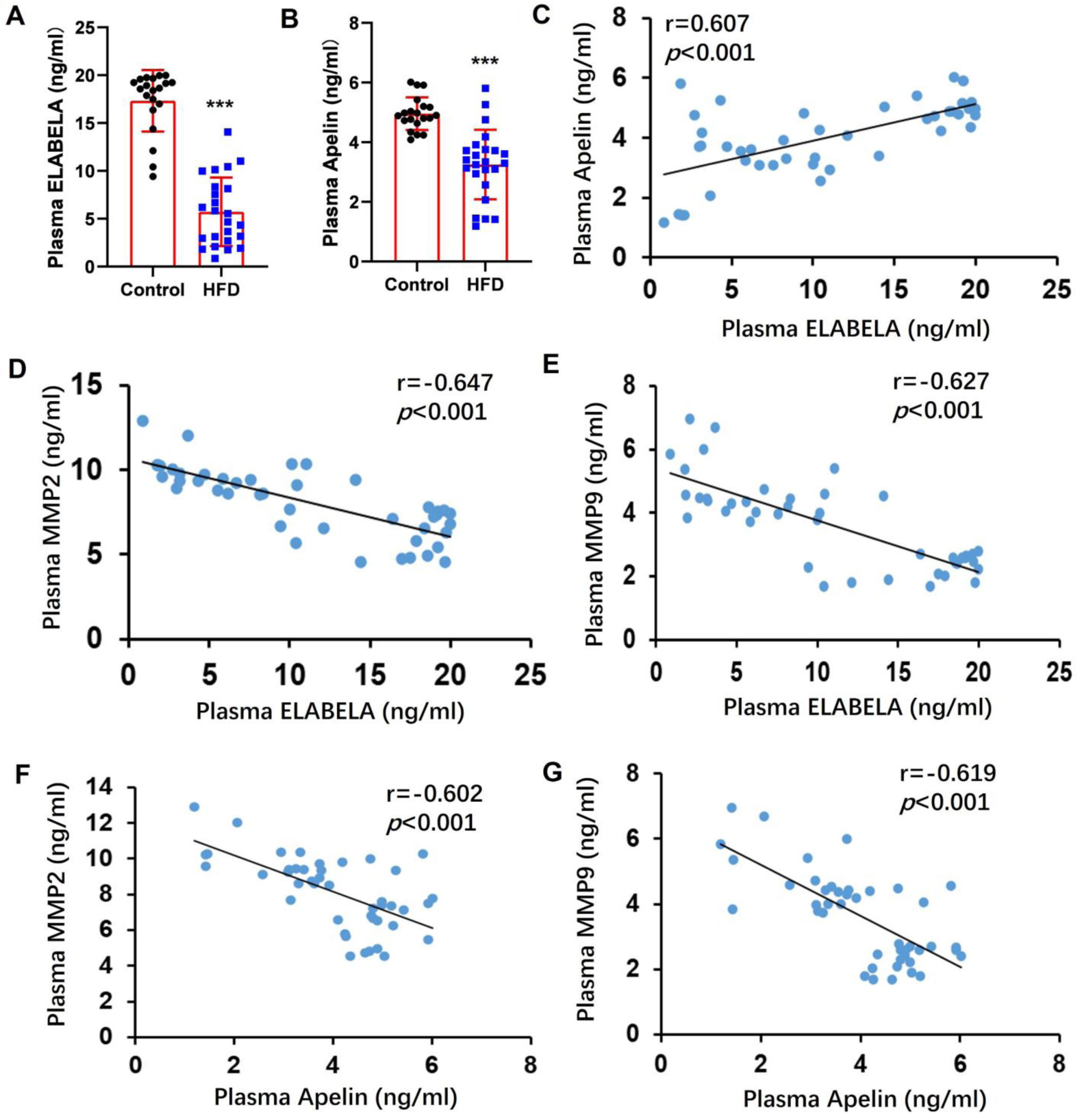
Plasma ELABELA and apelin levels in high-fat diet (HFD)-fed *ApoE*^-/-^ mice. (A and B) plasma ELABELA (A) and apelin (B) levels assessed by ELISA assays. n=20 for the Control group and n=25 for the HFD group. \*\*\**P*<0.001 *vs*. Control. (C) The correlation between plasma ELABELA and apelin. (D and E) The correlation between plasma ELABELA and matrix metalloproteinase 2 (MMP2) (D) and MMP9 (E). (F and G) The correlation between plasma apelin and MMP2 (F) and MMP9 (G).

### ELA-21 attenuated AS in *ApoE*^−/−^ mice

To study the impact of ELA-21 on AS, *ApoE*^−/−^ mice were fed a high-fat diet for 12 weeks, followed by a 4-week ELA-21 intervention (**Fig. 3A**). Compared to the HFD group, ELA-21 significantly reduced the lesion area, as reflected by the reduced lipid deposition assessed by Oil-Red-O staining (**Fig. 3B-C**) in full-length aortae. To further clarify the effect of ELA-21 on atherosclerotic plaques, Oil-red-O staining, H&E staining, and Masson’s trichrome staining were performed to evaluate lipid deposition, necrotic core size and plaque area, and collagen content within the aortic roots, respectively (**Fig. 3D**). There was a significant decrease in lipid deposition within the aortic root plaques in HFD+ELA-21 mice. The necrotic core size and plaque area within the aortic roots were significantly reduced in HFD+ELA-21 mice compared to HFD mice. Furthermore, the collagen fiber content within the aortic roots was also significantly increased in HFD mice with ELA-21 treatment. By immunofluorescence staining, although there was no difference in the macrophage content (CD68-positive cells) in the aortic root between the two groups, the α-SMA-positive area was significantly increased in the HFD+ELA-21 group (**Fig. 3E**). These results collectively indicate a significant therapeutic action of ELA-21 on AS by preventing the progression of atherosclerotic lesions and promoting a more stable plaque phenotype.

**Figure 3.**
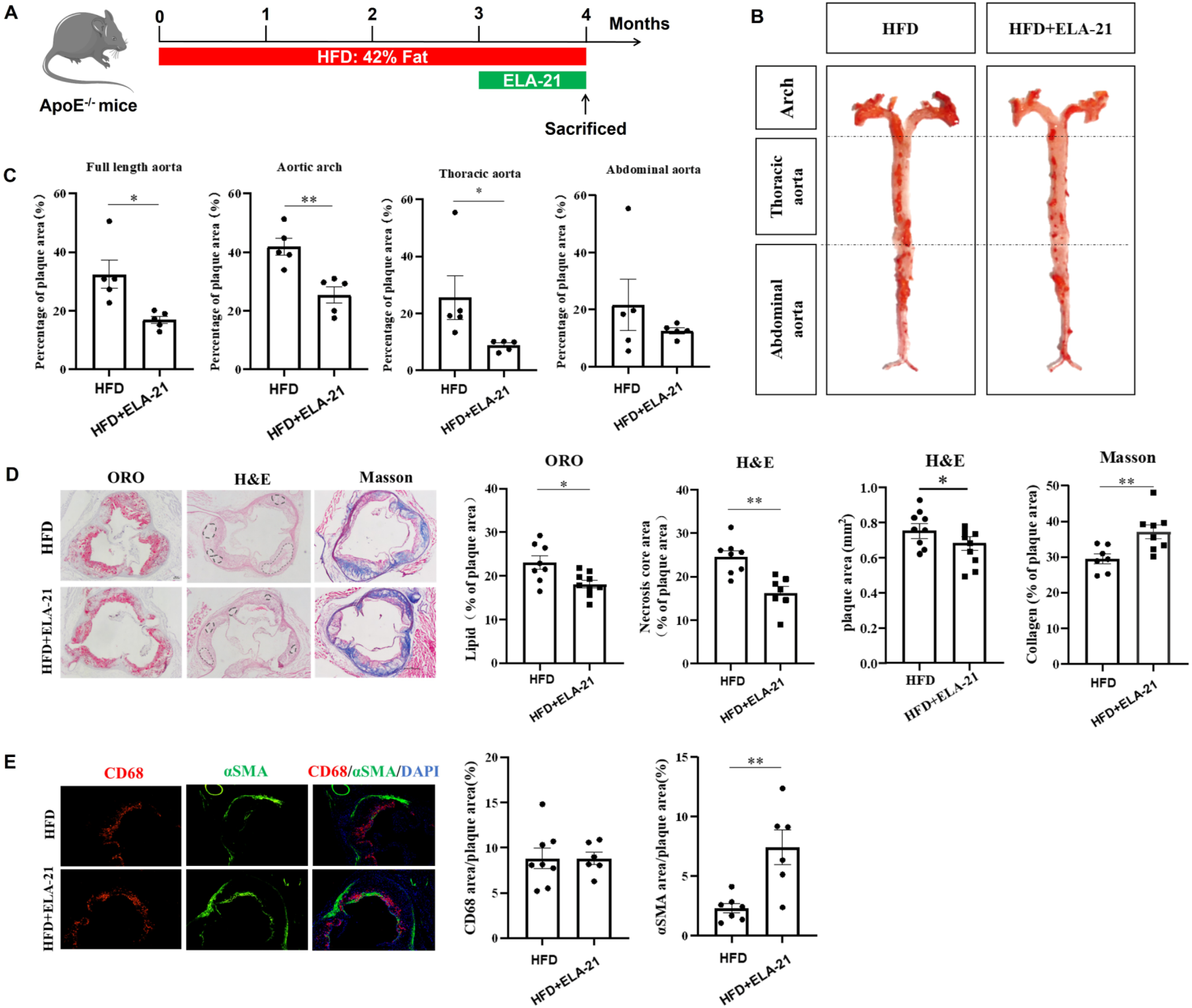
ELA-21 attenuated atherosclerosis in *ApoE*^−/−^ mice induced by a high-fat diet (HFD). (A) Experimental procedure for HFD-induced atherosclerosis in *ApoE*^−/−^ mice and ELABELA-21 (ELA-21) administration. (B) Oil-red-O staining of the full-length aorta from the *ApoE*^−/−^ mice. (C) Quantification of the plaques in the full-length aorta, aortic arch, thoracic aorta, and abdominal aorta. (D) Masson, Oil red O, and H&E staining of the aortic root from the *ApoE*^−/−^ mice, and the collagen fiber content, intra-plaque fat content, plaque area, and necrotic lesion area in the aortic root plaque were quantified. (E) CD68 and αSMA immunofluorescence staining in the aortic root from the *ApoE*^−/−^ mice (× 200) and the positive staining of CD68 and αSMA in the aortic root plaque was quantified. Data are mean ± SEM. \**P*<0.05, \*\**P*<0.01.

### ELA-21 suppressed inflammation and the formation of foam cells of macrophages

It is well-accepted that inflammation plays a significant role in the progression of AS, especially the process of plaque rupture^[1]^. We detected the mRNA expression levels of inflammatory cytokines in the full-length aortas and peritoneal macrophages from the *ApoE*^−/−^ mice by RT-qPCR. ELA-21 treatment significantly reduced aortic *Tnf-α*, *Il-1β*, *Il-12b*, *Inos*, *Il-6*, and *Mcp-1* mRNA expression in HFD-fed *ApoE*^−/−^ mice (**Figure 4A**). Macrophages, which can polarize towards pro-inflammatory M1 or anti-inflammatory M2 phenotypes, play a central role in atherosclerotic lesions^[21]^. In this study, the macrophage infiltration within the plaque area in the aortic roots from HFD-fed *ApoE*^−/−^ mice, as reflected by CD68 immunofluorescence staining, was not affected by ELA-21 (**Figure 3E**). However, EAL-21 reduced the percentage of iNOS^+^CD68^+^ cells while increasing the portion of Arg1^+^CD68^+^ cells within the aortic root plaques as reflected by the co-labeling of anti-CD68 antibody with anti-iNOS antibody (**Figure 4B**) or anti-Arg1 antibody (**Figure 4C**). In peritoneal macrophages from HFD-fed *ApoE*^−/−^ mice, ELA-21 significantly reduced the mRNA expression of *Il-1β*, *Tnf-α*, *Inos*, and *Il-12β* (the markers of M1 macrophages) while elevating the mRNA levels of *Fizz1*, *Il-10*, and *Cd206* (the markers of M2 macrophages) (**Figure 4D**). Thus, ELA-21 treatment modulated the balance between M1 and M2 macrophages toward a more anti-inflammatory status in HFD-fed *ApoE*^-/-^ mice. In addition, ELA-21 significantly decreased plasma levels of IL-1β, TNF-α, and IL-6 but increased plasma levels of IL-6 in HFD-fed *ApoE*^-/-^ mice (**Figure 4E**). To further clarify the effect of ELA-21 on M1 polarization, ELA-21 intervention was employed in LPS/IFN-γ-treated THP-1 and RAW264.7 macrophages *in vitro,* and RT-qPCR analysis was performed to detect the specific markers of M1 macrophages. In RAW264.7 cells, LPS/IFN-γ-stimulated *Tnf-α*, *Il-1β*, *Inos*, and *Il-12b* mRNA expression was consistently attenuated by ELA-21 treatment at least in high-dose (5 μM) (**Figure 5A**). The same experiments that were performed using the THP-1 cells yielded similar results. Administration of ELA-21 significantly abolished LPS/IFN-γ-induced upregulation of *TNF-α*, *IL-1β*, *IL-6*, *iNOS*, and *IL-12b* mRNA levels (**Figure 5B**). Thus, these results consistently suggest the inhibitory effect of ELA-21 on the M1 polarization of macrophages, at least in part contributing to the improved atherosclerotic lesions. To further verify whether the anti-inflammatory action of ELA-21 depends on the APJ in the macrophages, an APJ-specific inhibitor ML221 was employed. Interestingly, ELA-21-downregulated *TNF-α* and *IL-6* mRNA expression in THP-1 cells was further decreased by ML221 treatment (**Figure 5C**). These results suggest that the inhibitory effect of ELA-21 on macrophage inflammation may depend on an unknown receptor independent of the APJ.

**Figure 4.**
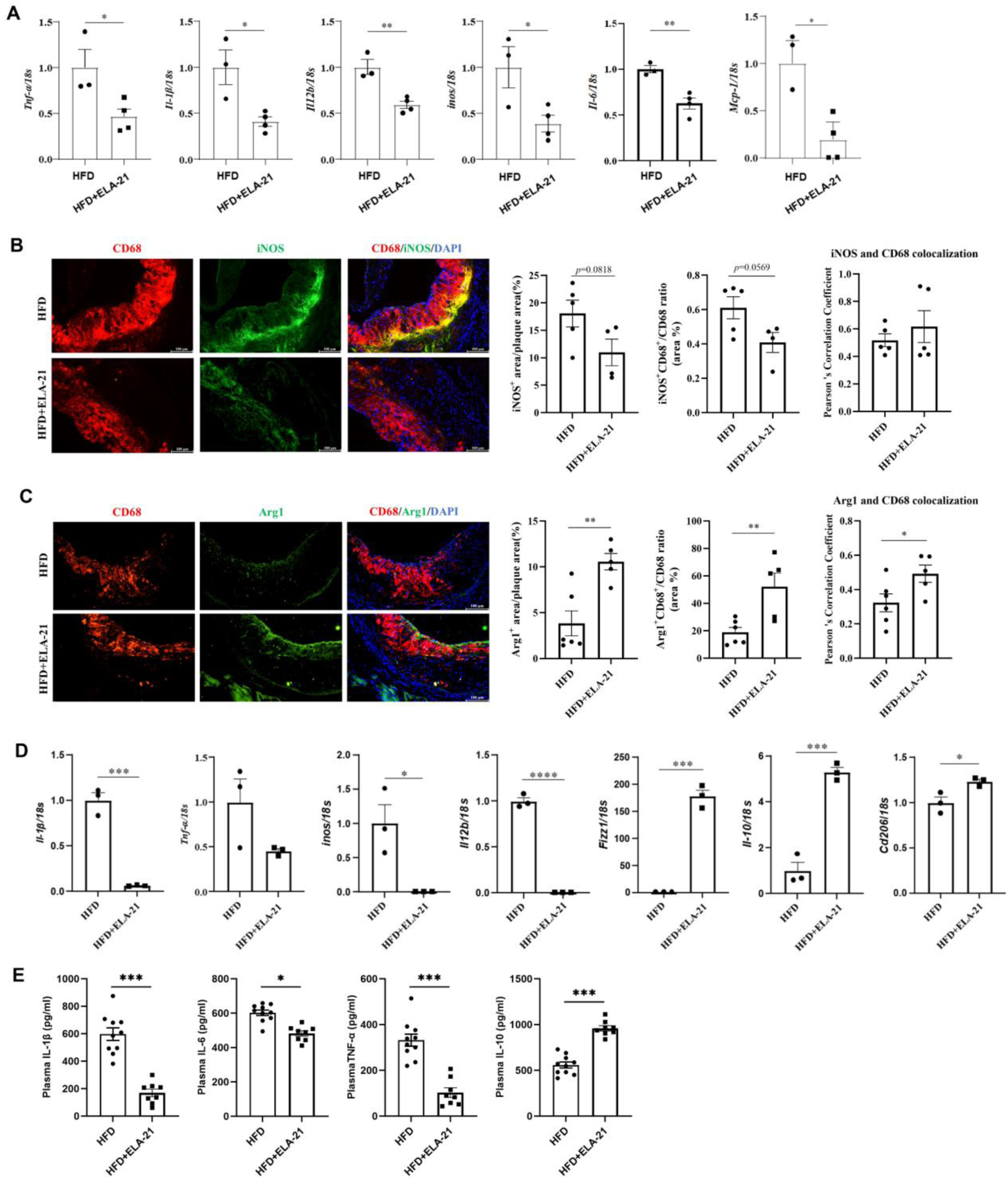
ELA-21 suppressed inflammation in *ApoE*^−/−^ mice induced by a high-fat diet (HFD). (A) RT-qPCR represents the expression of inflammatory factors in the full-length aorta from the *ApoE*^−/−^ mice. (B and C) iNOS (B) and Arg1 (C) immunofluorescence staining in the aortic root from the *ApoE*^−/−^ mice (×200) and he positive staining of iNOS and Arg1 in CD68-positive area was quantified. (D) RT-qPCR represents the expression of inflammatory factors in peritoneal macrophages from the *ApoE*^−/−^ mice. (E) Plasma levels of inflammatory factors in the *ApoE*^−/−^ mice assessed by ELISA. Data are mean ± SEM. \**P* < 0.05, \*\**P* < 0.01, \*\*\**P* < 0.001, \*\*\*\**P* < 0.0001.

**Figure 5.**
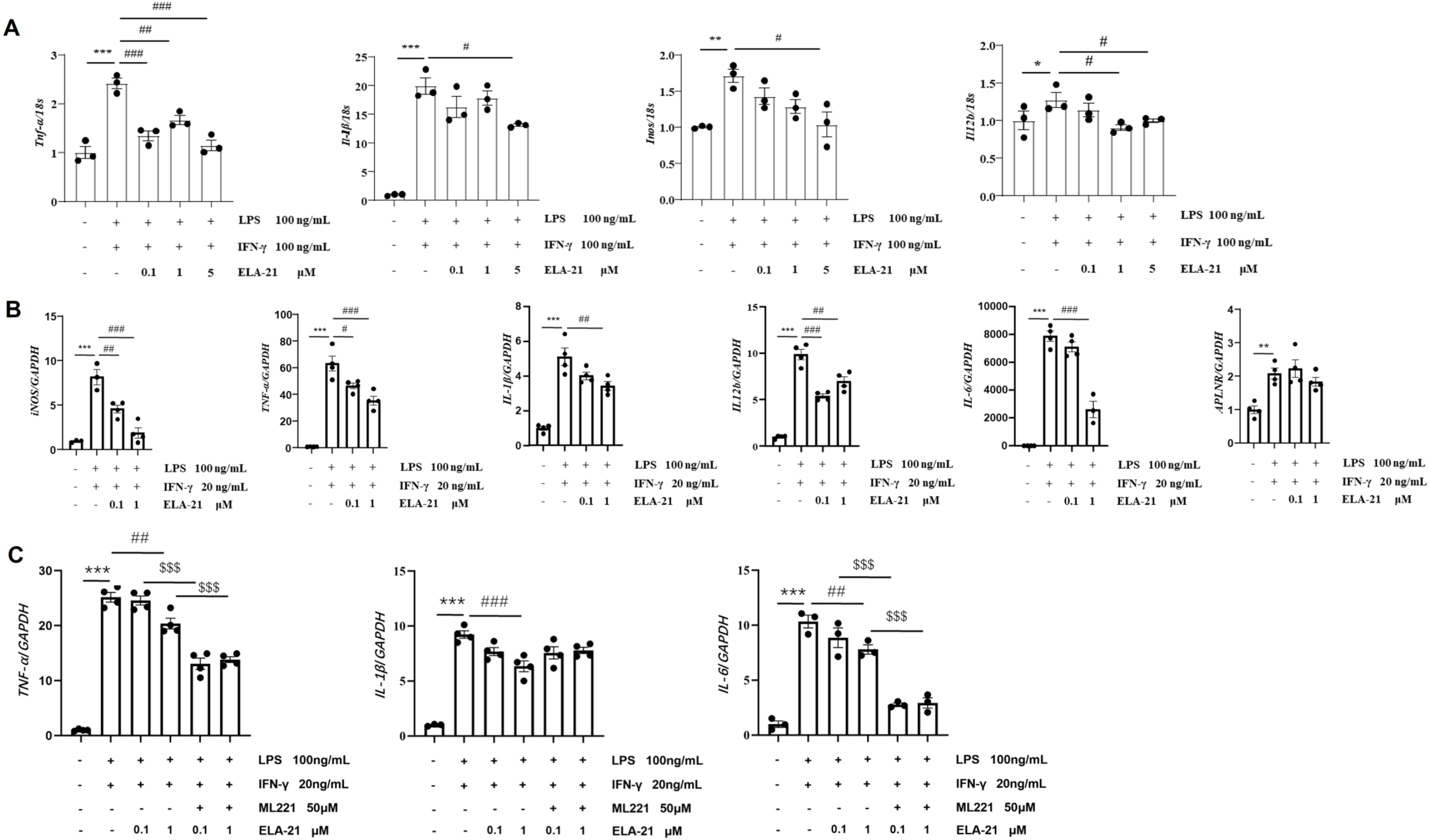
ELA-21 abolished the M1 polarization of macrophages *in vitro*. Mouse RAW264.7 (A) and human THP-1 (B and C) macrophages were stimulated by LPS/IFN-γ for 24 hours to induce M1 polarization in the presence or absence of ELA-21 or an APJ inhibitor ML221, RT-qPCR was performed to detect the mRNA levels of M1 polarization-related markers in macrophages. \**P* < 0.05, \*\**P* < 0.01, \*\*\**P* < 0.001, ^#^*P* < 0.05, ^##^*P* < 0.01, ^###^*P* < 0.001, ^$$$^*P*<0.001.

Macrophage foaming, one of the main hallmarks of early AS lesions, is caused by the accumulation of cholesterol esters due to the excessive uptake of oxidized low-density lipoprotein (ox-LDL)^[22]^. Foam cell formation and accumulation are hallmarks of AS progression and are new targets for fighting AS^[22]^. Here, we also investigated the possible impact of ELA-21 on macrophage foam cell formation *in vivo* and *in vitro*. First, HFD-caused lipid deposition in peritoneal macrophages from the *ApoE*^−/−^ mice assessed by Oil-Red-O staining was significantly abolished by ELA-21 administration (**Figure S1A**). Second, ELA-21 significantly attenuated ox-LDL incubation-induced lipid deposition in cultured THP-1 macrophages assessed by Oil-Red-O staining (**Figure S1B**). These results indicate an inhibitory action of ELA-21 on macrophage foaming, which may also contribute to its anti-atherosclerotic effect.

### ELA-21 enhanced Timp4 expression and reduced MMPs levels

To further clarify the potential mechanisms of ELA-21’s anti-atherosclerotic action, transcriptome sequencing of full-length aorta from HFD-fed *ApoE*^-/-^ mice was performed. As shown in **Figure 6A-C**, 160 genes were upregulated, and 153 genes were downregulated by ELA-21 administration. Among them, the five genes with the most significant increased expression levels include *Fkbp5*, *Timp4*, *Acer2*, *Rbp7*, and *Zbtb16*, while the five genes with the most significant decreased levels include *Has1*, *Sumo2*, *Nr1d1*, *Bcl3*, and *Il-6* (**Figure 6B**). The regulated genes were significantly enriched in lipid and AS-related pathways assessed by GO and KEGG enrichment analysis (**Figure 6C**). The imbalance between MMPs and their inhibitors (TIMPs) leads to the imbalance of proteolytic activity and adverse extracellular matrix remodeling, closely related to the initiation, progression, and rupture of atherosclerotic plaques^[23, 24]^. We thus detected the levels of MMPs and TIMPs in HFD-fed *ApoE*^−/−^ mice to clarify whether ELA-21 plays an anti-atherosclerotic role by regulating the MMPs and TIMPs. By RT-qPCR, administration of ELA-21 significantly decreased *Timp1*, *Timp2*, *Timp3*, *Mmp2*, and *Mmp9* mRNA levels but enhanced *Timp4* mRNA expression in the full-length aorta from HFD-fed *ApoE*^−/−^ mice (**Figure 6D**). We also conducted immunofluorescence staining to examine the expression of MMP2 and MMP9 in the aortic roots. The sections were co-labeled with anti-CD68 antibody. The signal within the plaque from anti-MMP2 antibody (**Figure 6E**) and anti-MMP9 antibody (**Figure 6F**) was significantly weakened by ELA-21, indicating the reduced MMP2 and MMP9 protein expression in the aortic roots after ELA-21 treatment. Furthermore, we compared plasma MMP2 and MMP9 levels between HFD and HFD+ELA-21 groups by using ELISA. The plasma levels of MMP2 and MMP9 in HFD-fed *ApoE*^−/−^ mice were significantly reduced by ELA-21 administration (**Figure 6G**). To further clarify the regulation of ELA-21 on MMP2 and MMP9 expression, we also detected the mRNA levels of *MMP2* and *MMP9* in cultured THP-1 cells stimulated by LPS/IFN-γ, in which ELA-21 treatment significantly decreased both *MMP2* and *MMP9* mRNA levels (**Figure 6H**). Thus, ELA-21 may enhance the stability of plaques by stimulating TIMP4 expression and inhibiting the release of MMPs.

**Figure 6.**
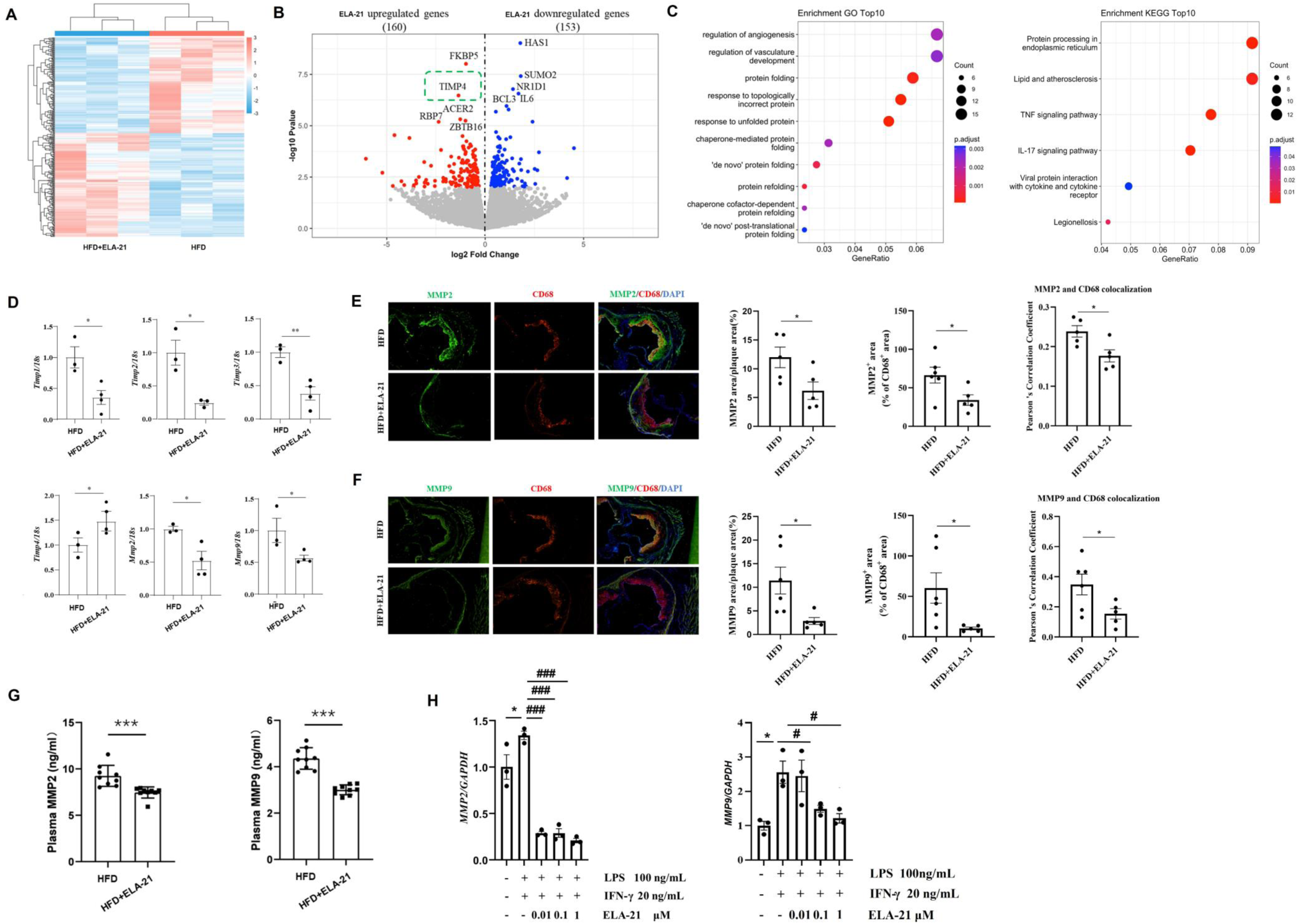
Regulation of ELA-21 on the expression of TIMPs and MMPs in the aorta from the high-fat diet (HFD)-fed *ApoE*^−/−^ mice. (A-C) mRNA sequencing results of the full-length aorta from the HFD-fed *ApoE*^−/−^ mice. (A) Cluster diagram of differentially expressed genes. (B) Volcanic diagram of differentially expressed genes. (C) GO and KEGG enrichment plot of differentially expressed genes. (D) RT-qPCR results of *Timps* and *Mmps* in the aorta from the HFD-fed *ApoE*^−/−^ mice. (E and F) MMP2 (E) and MMP9 (F) immunofluorescence staining in the aortic root from the HFD-fed *ApoE*^−/−^ mice (×200) and the positive staining of MMP2 and MMP9 in CD68-positive area was quantified. (G) Plasma MMP2 and MMP9 concentrations in the *ApoE*^−/−^ mice assessed by ELISA. (H) RT-qPCR results of *MMP2* and *MMP9* mRNA in the LPS/IFN-γ-treated THP-1 cells in the presence or absence of ELA-21. \**P* < 0.05, \*\**P* < 0.01, \*\*\**P* < 0.001, ^#^*P*<0.05, ^###^*P*<0.001.

### Modulation of ELA-21 on macrophage renin-angiotensin system

There is a close correlation between atherosclerotic plaque formation and the renin-angiotensin system (RAS), an important regulator of the inflammatory response ^[6, 25]^. To clarify the regulation of ELA-21 on the RAS in macrophages in the setting of AS, we conducted immunofluorescence staining to examine the expression of APJ, ACE, ACE2, and PRR in the aortic roots, which were co-labeled with anti-CD68 antibody. EAL-21 significantly increased the percentage of APJ^+^CD68^+^ (**Figure 7A**), ACE^+^CD68^+^ (**Figure 7B**), and ACE2^+^CD68^+^ (**Figure 7C**) cells while reducing the portion of PRR^+^CD68^+^ (**Figure 7D**) cells within the aortic root plaques. These results indicate the stimulatory role of ELA-21 on APJ, ACE, and ACE2 protein expression and the inhibitory effect of ELA-21 on PRR protein expression in macrophages under atherosclerotic conditions. Plasma sPRR levels in HFD-fed ApoE^−/−^ mice were significantly decreased by ELA-21 (**Figure 7E**). We also employed RT-qPCR to detect the mRNA levels of *Aplnr*, *Ace*, *Ace2*, and *Atp6ap2* in the full-length aorta from HFD-fed *ApoE*^−/−^ mice. As shown in **Figure 7F**, administration of ELA-21 significantly reduced *Ace* mRNA levels but elevated *Ace2* mRNA levels in the full-length aorta from HFD-fed *ApoE*^−/−^ mice, without affecting *Aplnr* and *Atp6ap2* mRNA levels. Furthermore, although LPS/IFN-γ stimulated both *ACE* and *ACE2* mRNA expression and reduced *ATP6AP2* mRNA levels, ELA-21 only further elevated *ACE2* mRNA levels in cultured THP-1 cells (**Figure 7F**). Interestingly, ML221 treatment further enhanced *ACE* and *ATP6AP2* mRNA expression but abolished ELA-21-upregulated *ACE2* mRNA levels (**Figure 7G**). Thus, the stimulation of ELA-21 on ACE2 expression may depend on the APJ, while the APJ may be required to inhibit ACE and PRR expression in macrophages.

**Figure 7.**
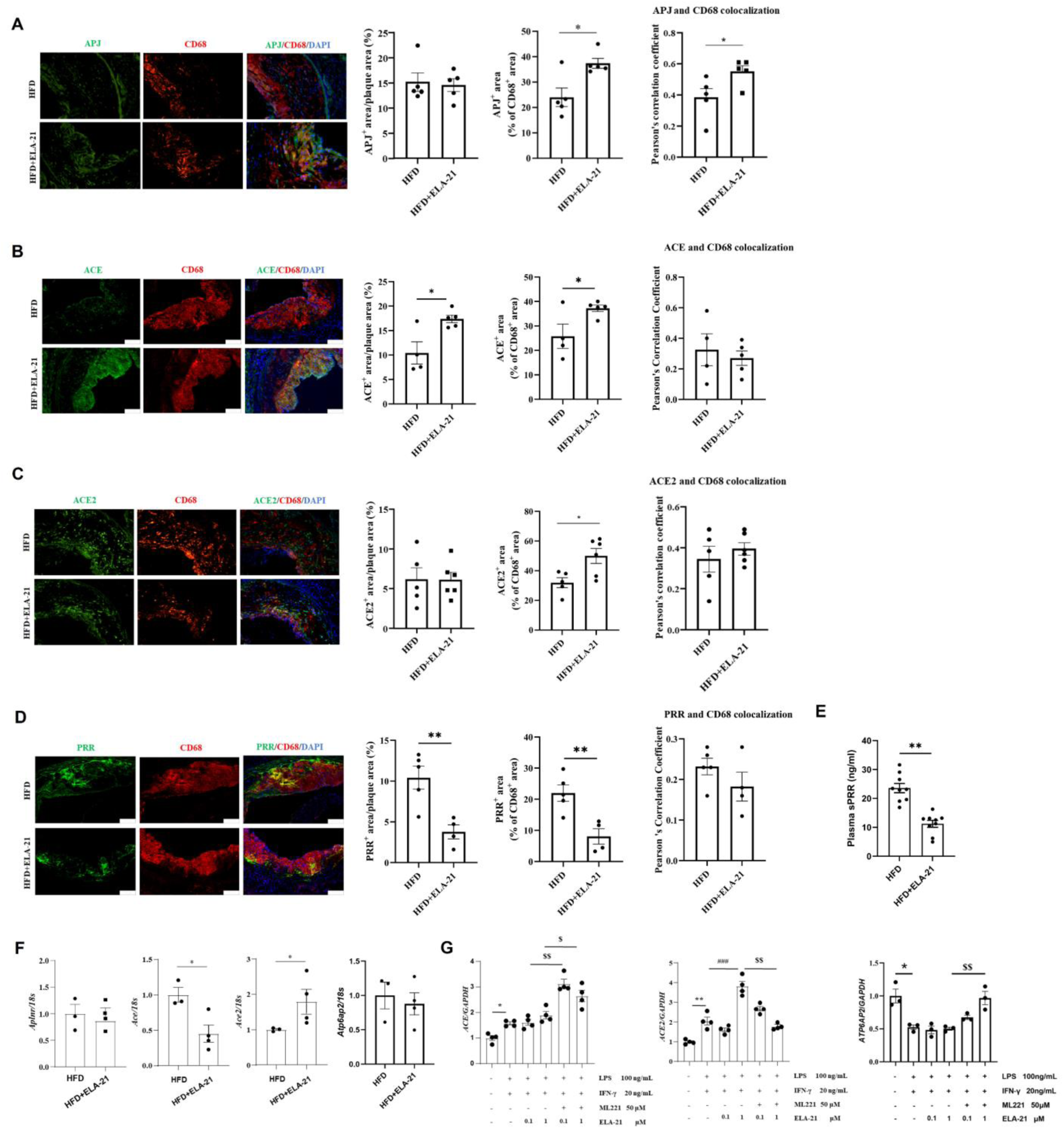
Regulation of ELA-21 on the expression of APJ, ACE, ACE2, and PRR in the aortic root from the high-fat diet (HFD)-fed *ApoE*^−/−^ mice. (A-D) APJ (A), ACE (B), ACE2 (C), and PRR (D) immunofluorescence staining in the aortic root from the HFD-fed *ApoE*^−/−^ mice (×200) and the positive staining of APJ, ACE, ACE2, and PRR in CD68-positive area was quantified. (E) Plasma sPRR concentrations in the HFD-fed *ApoE*^−/−^ mice assessed by ELISA. (F) RT-qPCR represents the expression of *Aplnr*, *Ace*, *Ace2*, and *Atp6ap2* mRNA in the full-length aorta from the HFD-fed *ApoE*^−/−^ mice. (G) RT-qPCR represents the expression of *ACE*, *ACE2*, and *ATP6AP2* mRNA in LPS/IFN-γ-treated THP-1 cells with and without ELA-21 or ML221 (an APJ inhibitor) treatment. \**P* < 0.05, \*\**P* < 0.01, ^###^*P*<0.001, ^$^*P* < 0.05, ^$$^*P* < 0.01.

## Discussion

The current study examined the potential correlation between ELABELA and AS progression. On the one hand, our data have demonstrated the diagnostic potential of ELABELA in AS, as reflected by the significant reduction of plasma ELABELA levels and its significant diagnostic value in AS patients. Plasma ELABELA levels negatively correlated with AS severity. On the other hand, our data have demonstrated the therapeutic potential of ELA-21 on AS, as reflected by the reduced lesion size within the aorta and enhanced stable atherosclerotic plaque phenotypes in HFD-fed *ApoE*^-/-^ mice with ELA-21 intervention. Mechanistically, the anti-atherosclerotic action of ELA-21 may be associated with the restored balance of the M1/M2 macrophage, enhanced expression of ACE and ACE2 in macrophages, and inhibited PRR system. Our findings may provide valuable insights for the use of ELABELA in AS patients and call for clinical evaluation of its anti-atherosclerotic efficacy and diagnostic effectiveness in individuals with atherosclerotic conditions.

In the present study, we systematically studied the potential relationship between ELABELA and AS and provided direct evidence for the potential diagnostic and therapeutic role of ELABELA in AS. First, in AS patients and atherosclerotic *ApoE*^−/−^ mice, plasma ELABELA levels were significantly decreased and inversely related to plasma MMP2 and MMP9, two important biomarkers of atherosclerotic plaque instability^[24]^, implying a possible certain association between plasma ELABELA levels and plaque vulnerability. Decreased plasma ELABELA levels may create a tendency towards vulnerable plaque. Second, plasma ELABELA exhibited a higher diagnostic value for AS than that of plasma apelin. ELABELA combined with MMP2 or MMP9 may significantly elevate the diagnostic accuracy of AS. Third, in HFD-fed *ApoE*^−/−^ mice, application of ELA-21 not only significantly reduced lipid accumulation in the aortas and atherosclerotic plaque size in aortic roots but also increased collagen accumulation and α-SMA expression in the aortic roots, accompanied with reduced MMP2 and MMP9 levels in the plasma and aortas and elevated aortic *Timp4* mRNA expression. Thus, ELA-21 not only reduced the lesion size but also enhanced atherosclerotic plaque stability. Interestingly, apelin-13 may only enhance atherosclerotic plaque stability without affecting the lesion size in HFD-induced atherosclerotic *ApoE*^−/−^ mice^[17]^. What’s more, APJ, the recognized receptor of ELABELA and Apelin, exhibits an opposite regulatory effect on AS to ELABELA and Apelin ^[18]^. These may indicate that the anti-atherosclerotic action of apelin-13 and ELA-21 is independent of the APJ. Indeed, APJ-independent actions of ELABELA via alternative unknown cell-surface receptors have already been reported in human embryonic stem cells^[26]^ and renal collecting duct cells^[27]^, in which APJ is lacking. Interestingly, although the levels of the total APJ protein in aortic roots and *Aplnr* mRNA in the aortas from HFD-fed *ApoE*^−/−^ mice were unchanged, the APJ levels in CD68-positive cells in plaques were significantly elevated upon ELA-21 treatment. This may imply the possible anti-atherosclerotic action of macrophage APJ, which may differ from that of APJ in other cell types like VSMCs. However, the elevated macrophage APJ may also be a compensatory response to inhibiting inflammatory signals downstream of the APJ. Thus, the effect of APJ in macrophages versus in VSMCs on AS progression needs further verification.

The M1 polarization of macrophage and subsequent foam cell formation and inflammatory response plays a central role in the AS^[28]^. Apelin/APJ signaling plays essential roles in macrophage growth, survival, and physiological and pathological functions^[29]^. In particular, apelin has been reported to inhibit macrophage inflammation^[15, 30, 31]^ and foam cell formation^[13, 14]^. Although several studies have demonstrated the anti-inflammatory effect of ELABELA in VSMCs^[32]^, ECs^[33]^, fibroblasts^[34]^, and peritoneal macrophages^[35]^, a report regarding the regulatory role of ELABELA on the polarization and foam cell formation of macrophages is still lacking. In the present study, ELA-21 treatment did not affect the macrophage infiltration, but significantly reduced iNOS^+^CD68^+^ macrophages and increased Arg1^+^CD68^+^ macrophages in the plaque in HFD-induced atherosclerotic *ApoE*^-/-^ mice. Besides, ELA-21 significantly decreased the mRNA expression of the markers of M1 macrophages and elevated the mRNA levels of the markers of M2 macrophages in peritoneal macrophages from these mice. These results consistently indicate the modulation of the balance between M1 and M2 macrophages toward a more anti-inflammatory status by ELA-21, which may contribute to the improvement of AS in HFD-fed *ApoE*^-/-^ mice. The inhibitory effect of ELA-21 on M1 polarization was further replicated *in vitro* cell experiments. In cultured THP-1 and RAW264.7 cells, LPS/IFN-γ-induced M1 polarization was significantly abolished by ELA-21 incubation. Interestingly, ELA-21-downregulated *IL-6* and *TNF-α* mRNA expression was further reduced by an APJ-specific inhibitor ML221 in LPS/IFN-γ-stimulated THP-1 cells. These results suggest that the inhibitory effect of ELA-21 on M1 polarization and inflammation may be independent of APJ, which may further support the APJ-independent anti-atherosclerotic action of ELA-21 via unknown receptors. Also, ELA-21 significantly attenuated the peritoneal macrophage foam cell formation from HFD-fed *ApoE*^-/-^ mice and ox-LDL-induced foam cell formation of THP-1 cells. Collectively, our data highlighted the anti-atherosclerotic effect of ELA-21 possibly by restoring the M1/M2 macrophage balance and inhibiting foam cell formation of macrophages.

The RAS, an important regulator of the inflammatory response, exhibits a close correlation with atherosclerotic plaque formation^[6, 25]^. ACE and ACE2 were present in both ECs, VSMCs, and macrophages in atherosclerotic lesions, but ACE was predominant in macrophages^[36, 37]^. Although the ACE inhibitor enalapril significantly inhibited AngI but not AngII-induced AS in *Ldlr*^-/-^ mice, the transplantation of macrophages lacking ACE into *Ldlr*^-/-^ mice has no effect on the atherosclerotic lesion in aortic roots and can only slightly reduce the lesion in aortic arch^[36]^. Similarly, vascular-specific deletion of ACE (ACE3/3 mice, only expressing ACE in liver and kidney)^[38]^ or transplantation of macrophages lacking ACE into *ApoE*^-/-^ mice^[39]^ does also not affect the formation of atherosclerotic plaques in mice. However, a series of reports from the Bernstein group have shown that macrophage-specific ACE overexpression (ACE10/10 mice) significantly reduced atherosclerotic plaque by enhancing macrophage lipid metabolism, cholesterol efflux, and efferocytosis in HFD-fed *ApoE*^-/-^ mice^[39]^ and AAV-PCSK9/HFD-induced atherosclerotic mice^[40, 41]^. Depart from the macrophage ACE, EC-specific deletion of ACE significantly reduced serum ACE activity without affecting the atherosclerotic lesion. In contrast, VSMC-specific deletion of ACE significantly inhibited the atherosclerotic lesion without affecting the serum ACE activity in HFD-fed *Ldlr*^-/-^ mice^[42]^. Thus, there may be a cellular difference in the regulation of ACE on AS that ACE in VSMCs rather than macrophages or ECs may be required for the occurrence and development of AS, but enhanced macrophage ACE may exhibit a significant anti-atherosclerotic action. In our study, although ELA-21 significantly reduced *Ace* mRNA expression in the aortas, it significantly enhanced ACE protein expression in CD68-positive cells in aortic roots from HFD-fed *ApoE*^-/-^ mice. However, although *ACE* mRNA expression in LPS/IFN-γ-stimulated THP-1 cells was unaffected by ELA-21, APJ inhibitor ML221 significantly elevated *ACE* mRNA levels. These results may indicate the potential inhibitory effect of APJ on *ACE* transcription, as compensatory negative feedback of the elevated ACE protein levels in macrophages. Overall, enhancement of macrophage ACE protein may contribute to the anti-atherosclerotic effect of ELA-21 by enhancing macrophage lipid metabolism and cholesterol efflux.

Compared with the diverse effects of ACE on AS, the role of ACE2 is much clearer and more consistent. Systemic ACE2 deletion significantly increased atherosclerotic plaques in *ApoE*^-/-^ mice, accompanied by significantly increased expression of adhesion factors, inflammatory cytokines, and MMP9, and the proinflammatory response in macrophages, ECs and VSMCs^[43, 44]^. Similarly, in HFD-fed *Ldlr*^-/-^ mice, ACE2 deletion in macrophages significantly increased atherosclerotic plaque and promoted the release of inflammatory cytokines and the adhesion of monocytes to ECs^[45]^. On the contrary, overexpression of ACE2 in monocytes or ECs leads to reduced endothelial adhesion and migration and the expression of adhesion factors^[46, 47]^. Recombinant adenovirus-mediated ACE2 overexpression ACE2 significantly reduced atherosclerotic plaques in HFD-fed *ApoE*^-/-^ mice^[47]^ and enhanced atherosclerotic plaque stability in New Zealand rabbits by improving endothelial function, inhibiting inflammatory response and monocyte adhesion, reducing macrophage infiltration, and inhibiting the ERK/p38, JAK/STAT, and NF-κB signaling pathways^[48]^. More interestingly, in *ApoE*^-/-^ mice with diabetes and *ApoE*^-/-^ mice lacking ACE2, recombinant mouse soluble ACE2 protein treatment significantly reduced atherosclerotic plaque^[49]^. In addition, Dimenazene, an activator of ACE2, significantly enhanced the stability of plaque by increasing the collagen content, reducing MMP9 expression and macrophage infiltration, and promoting M2 polarization of macrophages, accompanied by decreased expression of chemokines and inflammatory factors in plaques^[50]^. ACE2 overexpression inhibited M1 polarization of macrophages by suppressing the NF-κB signaling pathway^[51]^. These results consistently showed an anti-atherosclerotic role of ACE2 by inhibiting the formation and stability of atherosclerotic plaque. In the present study, administration of ELA-21 increased *Ace2* mRNA levels in the aortas and elevated ACE2 protein levels in CD68-positive cells in aortic roots from HFD-fed *ApoE*^-/-^ mice. In LPS/IFN-γ-stimulated THP-1 cells, *ACE2* mRNA expression was significantly enhanced by ELA-21, which was abolished by APJ inhibitor ML221. These results suggest the stimulation of ELA-21/APJ on ACE2 expression in both mRNA and protein levels in macrophages. Indeed, the protective action of ELABELA on cell injury by stimulating ACE2 expression has already been reported in cardiomyocytes^[52]^, aortic fibroblasts^[34]^, and renal tubular cells^[27]^. Overall, stimulation of macrophage ACE2 may contribute to the anti-atherosclerotic action of ELA-21 by inhibiting M1 polarization and subsequent inflammatory response.

The (pro)renin receptor (PRR) system, a key regulator of the RAS, exhibits a potential correlation with AS^[20, 53–56]^. Although the impact of PRR on AS may exhibit species differences^[54, 55]^, macrophage-specific PRR deletion significantly aggravated AS by augmenting inflammation and impairing cholesterol efflux in HFD-fed *ApoE*^-/-^ mice, caused by vacuolar H^+^-ATPase dysfunction rather than RAS activation ^[54]^. These results indicate the possible anti-atherosclerotic action of macrophage PRR by maintaining the activity of the vacuolar H^+^-ATPase. However, elevated circulating soluble PRR (sPRR) may contribute to AS progression through its pro-inflammatory effect by interacting and activating the angiotensin type 1 receptor (AT_1_R)^[57]^ in macrophages. In this regard, both *At_1_r* mRNA level in macrophages from the abdominal cavity^[58]^ and plasma sPRR level^[20, 59, 60]^ were elevated in mice with HFD feeding. AT_1_R inhibitor valsartan significantly reduced ox-LDL and *Il-1β* and *Tnf-α* mRNA levels in macrophages^[58]^. What’s more, elevated circulating soluble PRR (sPRR) levels were inversely correlated with ankle‒brachial index (ABI, an indicator of severe AS of the lower limbs) in hemodialysis patients^[53]^ and positively correlated with MMP2/MMP9 levels in atherosclerotic patients ^[20]^. These results imply the potential impact of sPRR on AS progression and atherosclerotic plaque stability. In this way, elevated plasma sPRR levels may lead to vulnerable plaque. Similar to our previous report on the inhibitory effect of ELA-32 on the PRR system^[27]^, here we found a reduced PRR protein level in CD68-positive cells within aortic roots and plasma sPRR concentrations in HFD-fed *ApoE*^-/-^ mice with ELA-21 treatment. Therefore, the anti-atherosclerotic role of ELA-21 may be related to the reduction of sPRR generation, which may further block AT_1_R-mediated macrophage inflammation and foaming, awaiting further experimental clarification.

We acknowledged that the present study has several shortcomings. On the one hand, the cohort study was a single-center observational study with a relatively small case number and lacking a healthy control group. A significant age difference between the AS and non-AS groups was observed. Age might influence the plasma ELABELA levels. The correlation of ELABELA with the severity of AS should be strengthened by adding other markers, such as the patients’ ABI. Future large-scale and multicenter studies and longitudinal follow-up studies are recommended to clarify the relationship between plasma ELABELA levels and the diagnosis, prognosis, mortality, and morbidity of AS patients. On the other hand, the specific receptor mediating the anti-atherosclerotic role of ELA-21 and the impact of macrophage APJ on AS is still unknown. The reasons for the simultaneous stimulation of ELA-21 on the protein expression of macrophage ACE and ACE2 within plaques are unclear since ELABELA has always been considered to inhibit ACE and PRR and stimulate ACE2 in cardiovascular tissues. Future studies are also needed to determine the relative contribution of the elevated macrophage ACE and ACE2 to the anti-atherosclerotic actions of ELA-21.

Nevertheless, our study, for the first time, showed the diagnostic and therapeutic potential of ELABELA in AS. ELABELA may be an effective diagnostic biomarker and a novel candidate therapeutic target for treating AS.

## DATA AVAILABILITY STATEMENT

The raw data supporting the conclusions of this article will be made available by Dr. Chuanming Xu without undue reservation.

## ETHICS STATEMENT

The studies involving human participants were reviewed and approved by the Affiliated Hospital of the Jiangxi University of Chinese Medicine (JZFYLL20230208002). The patients/participants provided written informed consent to participate in this study. The studies involving animals were reviewed and approved by the Animal Care and Use Committee approved the animal protocols at Jiangxi University of Chinese Medicine (No. JZLLSC20230254).

## Authors’ Contributions

Conceptualization, C.X.; methodology, L.T., X,Y., H.Y., S.S, W.T., J.X., C.L., Y. Z., M.W., M.Z., and L.Z.; validation, L.T., X,Y., H.Y., S.S, W.T., J.X., C.L., Y. Z., M.W., M.Z., and L.Z.; formal analysis and interpretation, L.T., X,Y., H.Y., S.S, W.T., J.X., C.L., Y. Z., M.W., M.Z., and L.Z.; investigation, C.X., L.T., X,Y., H.Y., S.S, W.T., J.X., C.L., Y. Z., M.W., M.Z., and L.Z.; data curation, L.T., X,Y., H.Y., S.S, W.T., J.X., C.L., Y. Z., M.W., M.Z., and L.Z.; writing-original draft preparation, C.X. and L.T.; writing-review and editing, C.X. and J.Y.; supervision, C.X.; project administration, C.X.; funding acquisition, C.X. All authors have read and approved the manuscript.

## Funding

This work was supported by grants from the National Natural Science Foundation of China (No. 82160051 and 32100908), the Jiangxi Provincial Natural Science Foundation (No. 20232BAB206018), the science and technology research project in Education Department of Jiangxi Province (No. GJJ2200904), Jiangxi “Double Thousand Plan” (No. jxsq2020101074), the Ph.D. Start-up Research Fund in Jiangxi University of Chinese Medicine (No. 2020BSZR009), the science and technology research project in Health Commission of Jiangxi Province (No. 202311143), Discipline of Chinese and Western Integrative Medicine in Jiangxi University of Chinese Medicine (Top Discipline of Jiangxi Province, No. zxyylxk20220103), the Scientific and Technological Innovation Team grant of the Jiangxi University of Chinese Medicine (No. CXTD22014), and the Jiangxi Key Laboratory grant in Science and Technology Department of Jiangxi Province (No.20202BCD42014).

## Consent for publication

Not applicable.

## Declaration of Competing Interest

The authors have no conflict of interest to disclose.

## Acknowledgments

Not applicable.

## Notes

### Competing Interest Statement

The authors have declared no competing interest.

### Clinical Trial

The studies involving human participants were reviewed and approved by the Affiliated Hospital of the Jiangxi University of Chinese Medicine (JZFYLL20230208002). We did not register the trial ID since we only collected blood samples from the peripheral veins of all patients.

### Author Declarations

The Ethics Committee of the Affiliated Hospital of the Jiangxi University of Chinese Medicine

